# Six-month antibody response to SARS-CoV-2 in healthcare workers assessed by virus neutralisation and commercial assays

**DOI:** 10.1101/2020.12.08.20245811

**Authors:** Antonin Bal, Mary-Anne Trabaud, Jean-Baptiste Fassier, Muriel Rabilloud, Kahina Saker, Carole Langlois-Jacques, Nicolas Guibert, Constance d’Aubarede, Adèle Paul, Dulce Alfaiate, Amélie Massardier-Pilonchery, Virginie Pitiot, Florence Morfin-Sherpa, Bruno Lina, Bruno Pozzetto, Sophie Trouillet-Assant, on behalf the COVID SER STUDY GROUP

## Abstract

We conducted a prospective study in healthcare workers (n=296) of the University Hospital of Lyon, France. Serum samples (n=296) collected six months after disease onset were tested using three commercial assays: the Wantai Ab assay detecting total antibodies against the receptor binding domain (RBD) of the S protein, the bioMerieux Vidas assay detecting IgG to the RBD and the Abbott Architect assay detecting IgG to the N protein. The neutralising antibody (NAb) titre was also determined for all samples with a virus neutralisation assay (VNA) using live virus. The positivity rate was 100% with the Wantai assay, 84.8% with the bioMerieux assay and 55.4% with the Abbott assay. Only 51% of HCWs were positive for the presence of NAb. Less than 10 % of HCWs had a NAb titre greater than 80. At a neutralising titre of 80, the area under the curves [IC 95%] was 0.71 [0.62-0.81], 0.75 [0.65-0.85] and 0.95 [0.92-0.97] for Wantai, Abbott and Vidas respectively. The data presented herein suggest that commercial assays detecting antibodies against the N protein must not be used in long-term seroprevalence surveys while the Wantai assay could be useful for this purpose. VNA should remain the gold standard to assess the protective antibody response, but some commercial assays could be used as first-line screening of long-term presence of NAb.

---

To the Editor,

Since the SARS-CoV-2 emergence in December 2019, one of the major concerns is the duration of immune protection after a first episode. This question is of paramount importance for healthcare workers (HCWs) who are a highly exposed population and among the first targets of vaccination programmes. To date, the persistence of SARS-CoV-2 antibodies in HCWs six months after disease onset (ADO) has not been studied with both a virus neutralisation test and commercial assays.

HCWs who experienced COVID-19 during the early phase of the pandemic were included in a prospective study conducted at the University Hospital of Lyon, France [1]. Serum samples collected six months ADO were tested using three commercial assays: the Wantai Ab assay that detects total antibodies against the receptor binding domain (RBD) of the S protein, the bioMérieux Vidas assay that detects IgG to the RBD, and the Abbott Architect assay that detects IgG to the N protein. The neutralising antibody (NAb) titre was determined by a virus neutralisation assay (VNA) using live virus as previously described [2].

A total of 296 HCWs were included; the median [interquartile range, IQR] age was 41 [32-51] years and 17.2% (51/296) were male. The median duration between symptom onset and inclusion was 186 [180-196] days. Of note, 8/296 HCWs (2.7%) were asymptomatic and the onset of disease was established on the basis of the median date of the RT-PCR positive result of the ward cluster. All participants were tested positive for SARS-CoV-2 serology at least two weeks after disease onset. The SARS-CoV-2 infection was also documented by RT-PCR test in 170 patients.

The positivity rate at six months ADO was 100% with the Wantai assay, 84.8% with the Vidas assay, and 55.4% with the Architect assay. Only 51% of HCWs were positive for the presence of NAb. Positive NAb titres ranged from 20 to 240. Only 27/296 (9.1%) had a NAb ≥ 80 (Figure 1A). No difference in positivity rates with any assay was observed between patients with a SARS-CoV-2 infection documented by RT-PCR and the rest of the cohort.

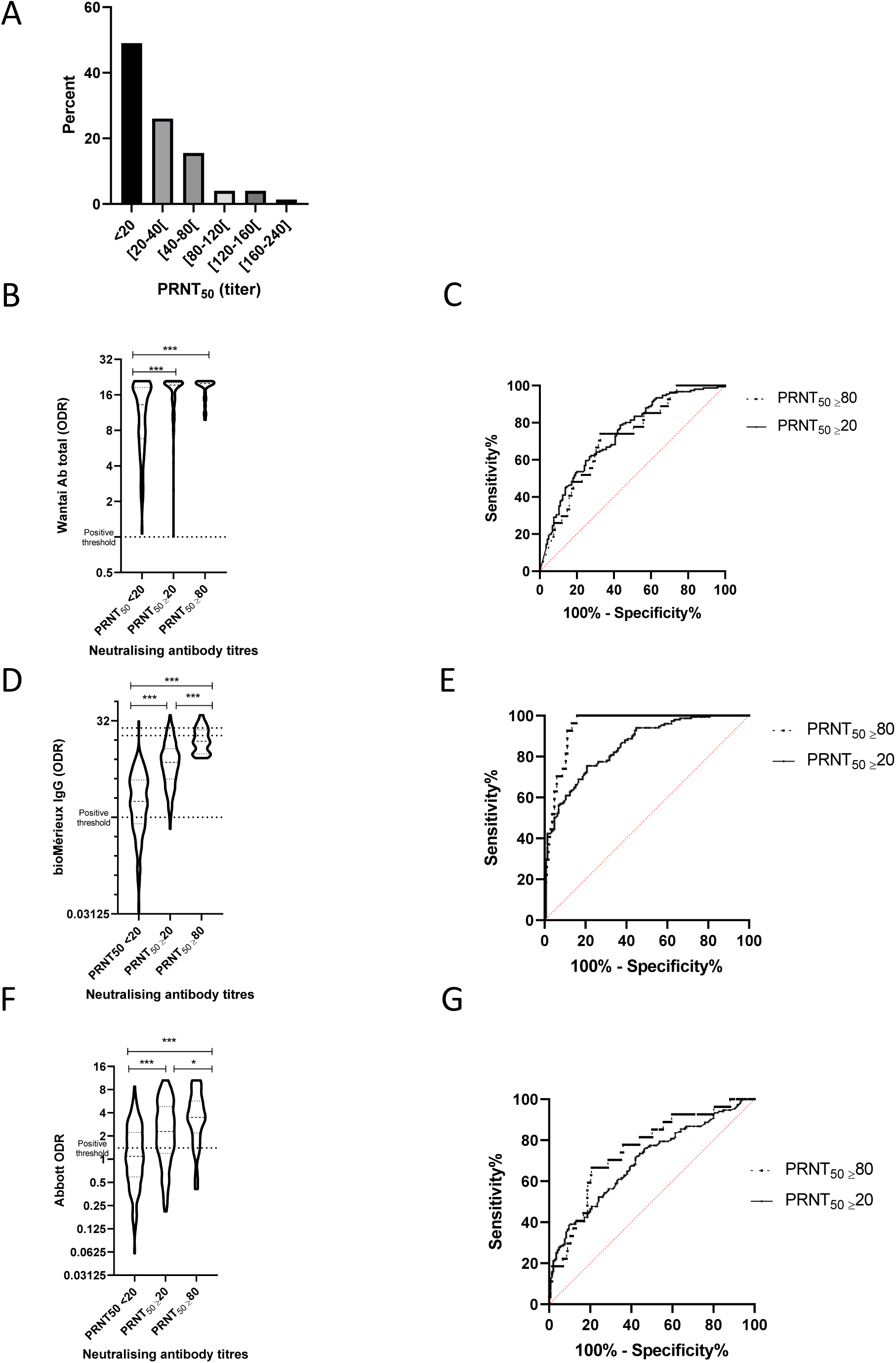
**A**. Distribution of neutralisation antibody titres in convalescent subjects (n=296) 6 months after SARS-CoV-2 infection. **B-D-F**. Violin plots describing ODR according to neutralising antibody titres. Dotted lines described positive threshold recommended by each manufacturer. Comparisons was performed using the Kruskal Wallis test followed by Dunn’s test. ***p<0.001, *p<0.05 **C-E-G**. ROC curves were built to estimate the performance of Wantai (C), bioMérieux (E) and Abbott (G) assays for detecting the presence of neutralising antibodies (PRNT_50_ ≥ 20-continuous line) and high neutralising antibody titre (PRNT_50_ ≥ 80-dotted line). ODR-Optical Density Ratio, PRNT-Plaque Reduction Neutralisation Titres.

Of the 296 HCWs, 6 (2.0%) developed a clinical form requiring hospitalisation; all were positive with the three serological assays and for the presence of NAb with a median titre of 40 (range: 30-160). By contrast, in asymptomatic HCWs, 8/8, 5/8, and 4/8 were positive with Wantai, Vidas, and Architect assays, respectively, and only 3/8 exhibited NAbs with low titres (range: 30-60).

The area under the ROC curve (AUC) was estimated for assessing the performance of serological assays for two NAb titres (PRNT_50_ ≥ 20 or PRNT_50_ ≥ 80; (Figure 1C, E, G). The highest AUCs were found with the Vidas assay: 0.85 (95% CI [0.81-0.89]) and 0.95 [0.92-0.97], respectively. The Wantai and Abbott assays had AUCs of, respectively, 0.73 [0.68-0.79] and 0.70 [0.64-0.76] for PRNT_50_ ≥ 20, and 0.71 [0.62-0.81], 0.75 [0.65-0.85] for PRNT_50_ ≥ 80. These results suggest that an optimised ratio with some commercial serological assay could be found to maximize the positive predictive value enabling to select individuals with a NAb titres ≥ 80. For instance, with the Vidas assay, the median [IQR] ratio for samples with PRNT_50_ ≥ 80 was 15.4 [9.7-22.7] vs 5.9 [3.3-9.2] for samples with a titre between 20 and 80 and 1.8 [0.8-3.8] for samples without NAb (Figure 1F). Among the 27 samples with NAb titre ≥ 80, all had a Vidas ratio above 8 compared to 31.5% and 3.5% of the samples with a titre between 20 and 80 or without NAb, respectively.

The findings of the present study indicate that, six months after infection, NAbs were no longer detected in about half of HCWs who presented mainly mild COVID-19. Overall, the detection of SARS-CoV-2 Abs with commercial tests was higher despite important heterogeneity between the assays evaluated herein. In a previous study [3], about 40% of asymptomatic subjects became negative for IgG to the N protein within 3 to 6 months, which is consistent with that presented herein for the Architect assay. This suggests that assays detecting only antibodies against the N protein must not be used in long-term seroprevalence surveys. By contrast, the Wantai assay could be very useful for epidemiological purposes as 100% of the HCWs were still positive at 6 months ADO. Although VNA should remain the gold standard to assess the protective antibody response, the data presented herein suggest that some commercial assays could be useful for first-line screening of long-term presence of NAb as previously reported within 4 months ADO [2,4].

Despite these observations on the decrease of NAbs in patients with mild COVID-19, it is important to note that they do not preclude the protective role of an anamnestic antibody response in previously exposed subjects, nor that of the long-term cellular immunity [5].

## Data Availability

All data are available in the manuscript.

## Ethics

Written informed consent was obtained from all participants; ethics approval was obtained from the national review board for biomedical research in April 2020 (Comité de Protection des Personnes Sud Méditerranée I, Marseille, France; ID RCB 2020-A00932-37), and the study was registered on ClinicalTrials.gov (NCT04341142).

## COVID-SER study group

Adnot Jérôme, Alfaiate Dulce, Bal Antonin, Bergeret Alain, Boibieux André, Bonnet Florent, Bourgeois Gaëlle, Brunel-Dalmas Florence, Caire Eurydice, Charbotel Barbara, Chiarello Pierre, Cotte Laurent, d’Aubarede Constance, Durupt François, Escuret Vanessa, Fascia Pascal, Fassier Jean-Baptiste, Fontaine Juliette, Gaillot-Durand Lucie, Gaymard Alexandre, Gillet Myriam, Godinot Matthieu, Gueyffier François, Guibert Nicolas, Josset Laurence, Lahousse Matthieu, Lina Bruno, Lozano Hélène, Makhloufi Djamila, Massardier-Pilonchéry Amélie, Milon Marie-Paule, Moll Frédéric, Morfin Florence, Narbey David, Nazare Julie-Anne, Oria Fatima, Paul Adèle, Perry Marielle, Pitiot Virginie, Prudent Mélanie, Rabilloud Muriel, Samperiz Audrey, Schlienger Isabelle, Simon Chantal, Trabaud Mary-Anne, Trouillet-Assant Sophie

## Acknowledgements

We thank all the personnel of the occupational health and medicine department of Hospices Civils de Lyon who contributed to the samples collection. Human biological samples and associated data were obtained from NeuroBioTec (CRB HCL, Lyon France, Biobank BB-0033-00046). We thank Karima Brahima and all members of the clinical research and innovation department for their reactivity (DRCI, Hospices Civils de Lyon). We thank Philip Robinson (DRCI, Hospices Civils de Lyon) for his help in manuscript preparation.

